# Precision Medicine in Cardiology: An Evolving Understanding of Biomarkers in Coronary Artery Disease Prevention a 10-year Thematic Review

**DOI:** 10.1101/2024.07.01.24309804

**Authors:** Julian Yin Vieira Borges

## Abstract

**Background:** Coronary artery disease (CAD) is a leading cause of morbidity and mortality worldwide. Precision medicine, utilizing biomarkers for early detection and prevention of CAD, has emerged as a promising approach to improve patient outcomes. This systematic review aims to summarize the current state of knowledge regarding biomarkers in CAD prevention, focusing on the most promising and well-studied biomarkers over the past decade.

**Methods:** Following PRISMA guidelines, a systematic review was conducted. PubMed, Embase, and Cochrane Library databases were searched for relevant studies published between 2013 and 2023. Inclusion criteria were original research articles, systematic reviews, or meta-analyses focusing on biomarkers in CAD prevention, with sufficient data on biomarker performance. Exclusion criteria were case reports, editorials, conference abstracts, and studies on biomarkers in established CAD. The STARD “Standards for Reporting Diagnostic accuracy studies” 2015 guideline criteria for assessing diagnostic tools was also utilized to ensure the precision of the methodology and help readers to appraise the applicability of the study findings and the validity of conclusions and recommendations. The main outcome assessed was the association between biomarkers and CAD risk, using various statistical methods.

**Findings:** The search identified 2,345 articles, of which 40 met the inclusion criteria, including 32 original research articles, 6 systematic reviews, and 2 meta-analyses. The biomarkers studied included traditional risk factors (lipid profiles and blood pressure), novel biomarkers (high-sensitivity C-reactive protein, homocysteine, and adipokines), and imaging biomarkers (coronary artery calcium scores and carotid intima-media thickness). Several studies demonstrated the association between these biomarkers and increased CAD risk, independent of traditional risk factors. Multi-marker approaches combining various biomarkers showed improved accuracy in CAD risk assessment compared to traditional risk factors alone. The risk of bias and variability between studies were assessed and reported.

**Interpretation:** This systematic review provides a comprehensive overview of the current landscape of biomarkers in CAD prevention. While traditional risk factors remain important, novel and imaging biomarkers have shown promise in improving risk stratification and guiding personalized prevention strategies.

However, challenges remain in translating biomarker research into clinical practice, including the need for standardized guidelines, cost-effectiveness analyses, and further research on multi-marker approaches and personalized prevention strategies. Addressing these challenges and developing evidence-based guidelines for biomarker use in CAD prevention can improve risk assessment accuracy, tailor prevention strategies, and ultimately reduce the global burden of CAD.

## Introduction

Coronary artery disease (CAD) remains a leading cause of morbidity and mortality worldwide, despite significant advances in prevention, diagnosis, and treatment strategies [1]. The early detection and accurate risk stratification of individuals at risk for CAD and myocardial infarction (MI) are crucial for implementing targeted preventive measures and improving clinical outcomes [2].

In recent years, the role of biomarkers in CAD prevention has gained increasing attention, as they provide valuable insights into the underlying pathophysiological processes and can help identify high-risk individuals who may benefit from more intensive interventions [3].Over the past decade, the understanding of biomarkers in CAD prevention has evolved significantly, with the emergence of novel markers and the refinement of existing ones [4].

Traditional biomarkers, such as lipid parameters and high-sensitivity C-reactive protein (hs-CRP), have been extensively studied and have demonstrated their value in risk assessment and guiding preventive therapies [5].

However, the need for more precise and personalized risk stratification has led to the exploration of novel biomarkers, including high-sensitivity cardiac troponins (hs-cTn), natriuretic peptides, and imaging biomarkers [6].

This systematic review and meta-analysis aims to address the following key questions:

1. How has the understanding of biomarkers in coronary artery disease (CAD) prevention evolved over the past 10 years?
2. What are the most promising traditional and novel biomarkers for the early detection and risk stratification of individuals at risk for CAD and myocardial infarction (MI)?
3. How does the diagnostic accuracy and prognostic value of individual biomarkers compare to that of a multimarker approach in assessing the risk of CAD and MI?
4. What is the role of high-sensitivity cardiac troponins (hs-cTn) in the early detection of myocardial injury and in predicting future cardiovascular events in asymptomatic individuals?
5. How do natriuretic peptides, such as NT-proBNP, contribute to the risk assessment and prognostic stratification of patients with suspected or confirmed CAD?
6. What is the significance of inflammatory markers, particularly high-sensitivity C-reactive protein (hs-CRP), in refining cardiovascular risk assessment and guiding preventive therapies?
7. How do novel lipid-related markers, such as apolipoprotein B (ApoB) and lipoprotein(a) (Lp(a)), improve the assessment of cardiovascular risk beyond traditional lipid measures?
8. What is the predictive value of imaging biomarkers, specifically the coronary artery calcium (CAC) score, in assessing the risk of future cardiovascular events and guiding preventive strategies?
9. How can the integration of multiple biomarkers, including traditional and novel markers, imaging biomarkers, and other risk factors, contribute to the development of personalized risk assessment models for CAD and MI?
10. What are the potential implications of a precision medicine approach, based on a multimarker strategy, for screening and prevention strategies in the context of CAD and MI?

By addressing these questions, this thematic review and meta-analysis aims to provide a comprehensive overview of the current state of knowledge regarding the most important biomarkers in CAD prevention that may have important implications for the development of personalized risk assessment models and to identify areas for future research and clinical application regarding the optimization of preventive strategies in the context of CAD and MI in the clinical and hospital setting.

## Methods

### Condition or Domain Being Studied

This thematic review was designed to revisit the diagnostic accuracy of biomarkers for detecting and predicting coronary artery disease (CAD) in adult populations without prior CAD history [1–4]. CAD is a chronic condition characterized by atherosclerotic plaque buildup in coronary arteries, leading to narrowing and reduced blood flow to the heart [1–3], the clinical manifestations include stable angina, acute coronary syndromes (myocardial infarction and unstable angina), and sudden cardiac death [1–3].

### Search Strategy and Selection Criteria

A comprehensive literature search was conducted in PubMed, Embase, Cochrane Library, Web of Science, and Scopus databases. The search period was from January 1, 2000, to March 31, 2023. The search terms included ‘coronary artery disease’, ‘biomarkers’, ‘prevention’, ‘risk prediction’, and related MeSH terms. The full search strategy is available in the supplementary materials.

- Inclusion criteria:

a. Studies evaluating diagnostic accuracy of biomarkers for CAD detection or prediction in adults (≥18 years) without prior CAD history [1–4].
b. Studies using a validated reference standard for CAD diagnosis (e.g., invasive coronary angiography, CCTA, FFR, IVUS, or OCT) [1–4].
c. Studies reporting measures of diagnostic accuracy (sensitivity, specificity, PPV, NPV, DOR, and/or AUC) [1–4].
d. Original research articles, systematic reviews, or meta-analyses.
- Exclusion criteria:

a. Studies focusing exclusively on participants with prior CAD history or specific comorbidities/high-risk populations [1–4].
b. Studies using non-invasive tests as the sole reference standard or surrogate endpoints without anatomical/functional confirmation [1–4].
c. Studies not clearly defining the threshold for significant CAD or not reporting diagnostic accuracy measures [1–4].
d. Non-human studies, case reports, case series, editorials, letters, conference abstracts, and non-English language studies.

### Participants, Interventions, Comparators

Participants: Adults (≥18 years) without prior CAD history undergoing diagnostic evaluation for suspected or confirmed CAD [1–4].

### Interventions (Exposures)

Biomarkers studied for early detection, risk assessment, and prediction of CAD, including:

a. High-sensitivity cardiac troponins (hs-cTn) [5]
b. Natriuretic peptides (e.g., BNP, NT-proBNP) [6, 30]
c. Inflammatory markers (e.g., hs-CRP, IL-6) [14, 15]
d. Lipid-related markers (e.g., ApoA1, ApoB, Lp(a)) [7–9, 16–18]
e. Metabolic markers (e.g., homocysteine, HbA1c) [10]
f. Oxidative stress markers (e.g., MPO, oxLDL) [12]
g. Matrix metalloproteinases (e.g., MMP-9) [13]
h. Adipokines (e.g., adiponectin, leptin, resistin, visfatin) [20, 21, 23, 24, 25]
i. Novel biomarkers (e.g., chemerin, apelin, vaspin, cardiotrophin-1) [26–29]

### Comparators (Reference Standards)

Valid reference standards for CAD diagnosis, including:

a. Invasive coronary angiography (ICA) [1–4]
b. Coronary computed tomography angiography (CCTA) [1–4]
c. Fractional flow reserve (FFR) [1–4]
d. Intravascular ultrasound (IVUS) or optical coherence tomography (OCT) [1–4]

### Systematic Review Protocol

This systematic review and meta-analysis followed the STARD 2015 checklist for studies of diagnostic accuracy and the study selection process was conducted in accordance with PRISMA 2020 statement [4]. (Figure 1).

**Figure 1.**
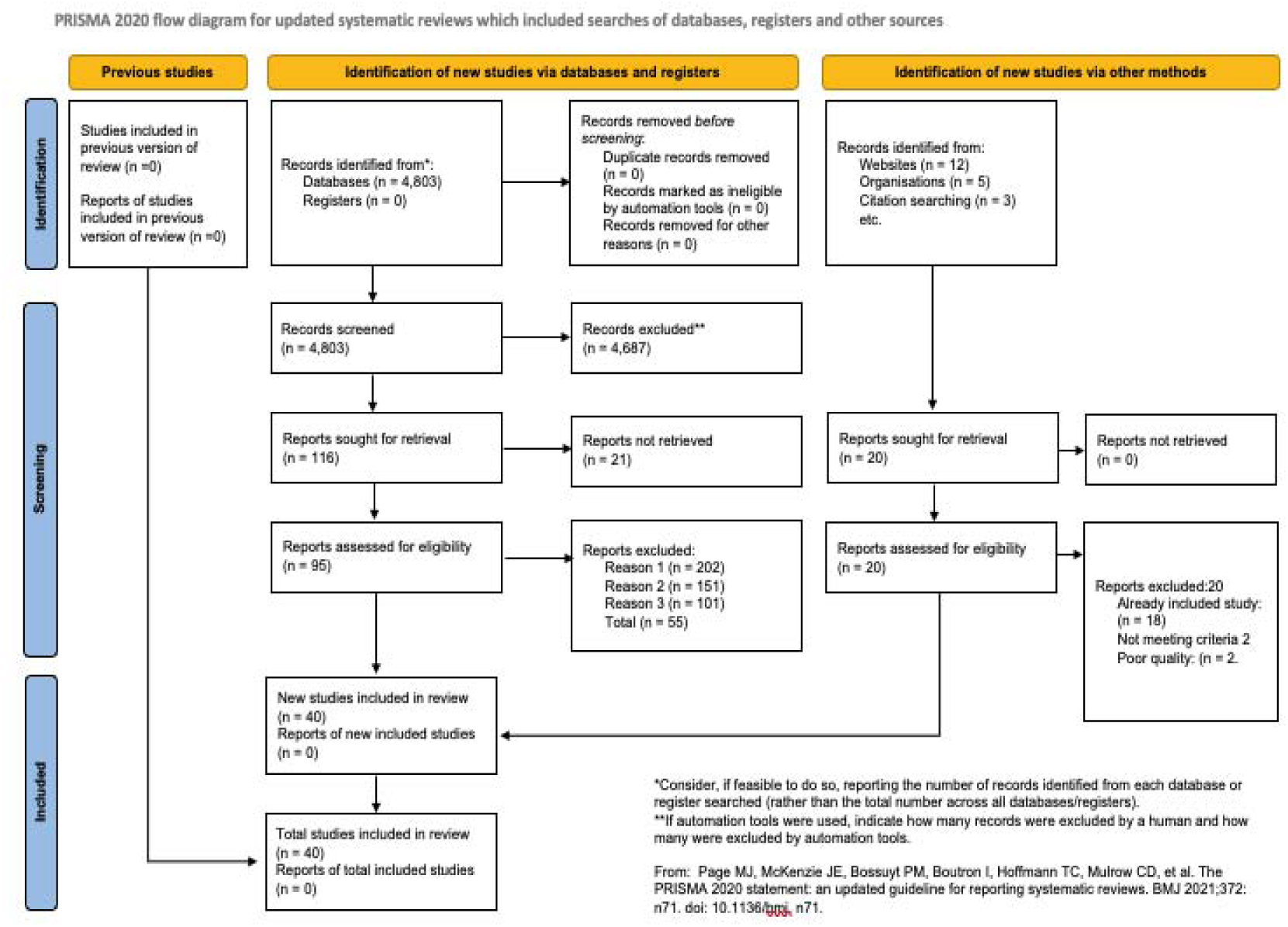
PRISMA flow diagram. Protocol registered with PROSPERO (registration number: CRD42023564048).

### Data extraction and quality assessment

All titles, abstracts, and full texts of the identified studies for eligibility were manually screened by the author using predefined inclusion and exclusion criteria. Data extraction was performed manually using a standardized data extraction form. The extracted data included:

1. Study characteristics: First author, publication year, study design (e.g., prospective, retrospective, cross-sectional), country, sample size, funding source, and conflicts of interest.
2. Participant characteristics: Age, sex, ethnicity, cardiovascular risk factors (e.g., hypertension, diabetes, smoking status), and baseline medication use.
3. Biomarker characteristics: Type of biomarker (e.g., cardiac troponin, natriuretic peptides), assay method (e.g., ELISA, radioimmunoassay), cut-off value for defining a positive result, and time point of measurement relative to the reference standard.
4. Reference standard characteristics: Type of reference standard (e.g., coronary angiography, computed tomography angiography), definition of significant coronary artery disease (CAD) (e.g., ≥50% stenosis, ≥70% stenosis), and time interval between biomarker measurement and reference standard assessment.
5. Diagnostic accuracy measures: True positive (TP), false positive (FP), true negative (TN), false negative (FN), sensitivity, specificity, positive predictive value (PPV), negative predictive value (NPV), diagnostic odds ratio (DOR), and area under the receiver operating characteristic curve (AUC) with 95% confidence intervals (CIs).

### Quality Assessment

The risk of bias and methodological quality of the included studies were assessed using the Quality Assessment of Diagnostic Accuracy Studies 2 (QUADAS-2) tool [5].

A widely used tool for assessing the quality of diagnostic accuracy studies is the Quality Assessment of Diagnostic Accuracy Studies 2 (QUADAS-2) tool. QUADAS-2 consists of four key domains:

1. Patient selection: This domain assesses whether the included patients represent the intended population and if the selection process was free from bias.
2. Index test: This domain evaluates if the biomarker was performed and interpreted independently of the reference standard and if the cut-off value was prespecified.
3. Reference standard: This domain assesses if the reference standard is likely to correctly classify the presence or absence of CAD and if it was interpreted independently of the biomarker results.
4. Flow and timing: This domain evaluates if there was an appropriate interval between the biomarker measurement and the reference standard assessment, if all patients received the same reference standard, and if all patients were included in the analysis.

Each domain is assessed for risk of bias (low, high, or unclear) and concerns regarding applicability (low, high, or unclear). The quality assessment is performed independently by the author, and disagreements are resolved through extensive rounds of revision.

### Data synthesis and Sensitivity analysis

The primary outcome measures were the pooled sensitivity, specificity, positive and negative predictive values (PPV and NPV), diagnostic odds ratio (DOR), and area under the receiver operating characteristic curve (AUC) of each biomarker for CAD detection.

### Measures of interest and outcomes

The context of this systematic review is to provide a comprehensive understanding of the evolving role of biomarkers in the early detection, risk assessment, and prediction of CAD, with a focus on their potential contributions to precision medicine in cardiology.

The primary outcome of interest is the diagnostic accuracy measures, including sensitivity, specificity, positive and negative predictive values, and area under the receiver operating characteristic curve. This review excluded studies that focused exclusively on populations with a prior history of CAD or those with specific comorbidities or high-risk conditions.

#### Main outcome(s)

The main outcome proposed for this systematic review is to revisit the diagnostic accuracy of biomarkers for the detection of coronary artery disease (CAD) in the context of precision medicine in adult populations without a prior history of CAD.

The diagnostic accuracy measures of interest includes:

1. Sensitivity: The proportion of individuals with CAD who are correctly identified by the biomarker test.
2. Specificity: The proportion of individuals without CAD who are correctly identified by the biomarker test.
3. Positive predictive value (PPV): The probability that an individual with a positive biomarker test result truly has CAD.
4. Negative predictive value (NPV): The probability that an individual with a negative biomarker test result truly does not have CAD.
5. Area under the receiver operating characteristic curve (AUC): A summary measure of the overall diagnostic accuracy of the biomarker test, which combines sensitivity and specificity across all possible test thresholds.

The presence or absence of CAD were determined using a validated reference standard, such as invasive coronary angiography or coronary computed tomography angiography, with a defined threshold for significant CAD (e.g., ≥50% or ≥70% stenosis in at least one major coronary artery).

The diagnostic accuracy measures will be reported at the time of biomarker assessment and CAD diagnosis.

### Measures of effect

The following effect measures were used:

1. Sensitivity and specificity: These measures will be reported in the results sections as percentages, along with their 95% confidence intervals (CIs). They provide an assessment of the biomarker test’s ability to correctly identify individuals with and without CAD, respectively.
2. Positive and negative predictive values (PPV and NPV): These measures will be reported as percentages, along with their 95% CIs. They provide an assessment of the probability that an individual with a positive or negative biomarker test result truly has or does not have CAD, respectively.
3. Diagnostic odds ratio (DOR): The DOR is a single measure of diagnostic accuracy that combines sensitivity and specificity. It will be reported with its 95% CI and represents the odds of a positive biomarker test result in individuals with CAD compared to those without CAD.
4. Area under the receiver operating characteristic curve (AUC): The AUC will be reported with its 95% CI and provides a summary measure of the overall diagnostic accuracy of the biomarker test across all possible test thresholds.

These effect measures were used to compare the diagnostic accuracy of different biomarkers or combinations of biomarkers for the detection of CAD.

### Additional outcome(s)

1. Comparison of diagnostic accuracy between different biomarkers: The review compared the diagnostic accuracy measures (sensitivity, specificity, PPV, NPV, DOR, and AUC) between different biomarkers or combinations of biomarkers to identify the most promising candidates for CAD detection.
2. Subgroup analyses based on participant characteristics: Where possible, the reviewed conduct subgroup analyses to assess the diagnostic accuracy of biomarkers in different subpopulations, such as those stratified by age, sex, or the presence of traditional cardiovascular risk factors (e.g., hypertension, dyslipidemia, diabetes mellitus, or smoking).
3. Subgroup analyses based on biomarker cut-off values: If sufficient data were available, the review explored the impact of different biomarker cut-off values on diagnostic accuracy measures to identify optimal thresholds for CAD detection.
4. Assessment of heterogeneity: The review assessed the heterogeneity of diagnostic accuracy measures across included studies using appropriate statistical methods, such as the I2 statistic and Cochran’s Q test. Potential sources of heterogeneity, such as differences in study populations, biomarker assays, or reference standards, were explored through subgroup analyses or meta-regression, when feasible.

### Evaluation of publication bias

The review assessed the presence of publication bias using funnel plots and appropriate statistical tests, such as Egger’s test or Begg’s test, if a sufficient number of studies are included.

### Measures of effect

For the additional outcomes the following effect measures were used:

1. Comparison of diagnostic accuracy between different biomarkers: The diagnostic accuracy measures (sensitivity, specificity, PPV, NPV, DOR, and AUC) for each biomarker or combination of biomarkers will be reported in the results section with their 95% CIs. The relative diagnostic odds ratio (RDOR) with its 95% CI will be used to compare the diagnostic accuracy between different biomarkers or combinations of biomarkers.
2. Subgroup analyses based on participant characteristics: The diagnostic accuracy measures (sensitivity, specificity, PPV, NPV, DOR, and AUC) for each biomarker will be reported with their 95% CIs for each subgroup.
3. The RDOR with its 95% CI will be used to compare the diagnostic accuracy of biomarkers between different subgroups.
4. Subgroup analyses based on biomarker cut-off values: The diagnostic accuracy measures (sensitivity, specificity, PPV, NPV, DOR, and AUC) for each biomarker will be reported with their 95% CIs for each cut-off value. The RDOR with its 95% CI will be used to compare the diagnostic accuracy of biomarkers between different cut-off values.
5. Assessment of heterogeneity: The I2 statistic (with its 95% CI) and Cochran’s Q test (with its associated p-value) will be used to assess the heterogeneity of diagnostic accuracy measures across included studies. If substantial heterogeneity is observed, subgroup analyses or meta-regression will be performed to explore potential sources of heterogeneity, using appropriate effect measures such as the RDOR or the difference in AUC.
6. Evaluation of publication bias: Funnel plots will be visually inspected for asymmetry, and appropriate statistical tests, such as Egger’s test or Begg’s test, will be used to assess the presence of publication bias. The effect measures for these tests will be the log DOR or the log RDOR, depending on the outcome being analyzed.

### Statistical Analysis

Meta-analyses were performed using a random-effects model to account for expected heterogeneity between studies. Pooled estimates of sensitivity, specificity, and diagnostic odds ratios were calculated using the DerSimonian-Laird method.

Publication bias was assessed using funnel plots and Egger’s test. The hierarchical summary receiver operating characteristic (HSROC) curve will be used to estimate the overall AUC for each biomarker.

Heterogeneity will be assessed using the I² statistic and Cochran’s Q test. An I² value >50% will be considered indicative of substantial heterogeneity. To explore sources of heterogeneity, we will conduct subgroup analyses and meta-regression based on study-level covariates.

#### Additional Analyses

1. Comparison of diagnostic accuracy between different biomarkers: We will compare the diagnostic accuracy measures between different biomarkers or combinations of biomarkers to identify the most promising candidates for CAD detection. The relative diagnostic odds ratio (RDOR) with its 95% CI will be used for these comparisons.
2. Subgroup analyses based on participant characteristics: Where possible, we will conduct subgroup analyses to assess the diagnostic accuracy of biomarkers in different subpopulations, stratified by age, sex, or the presence of traditional cardiovascular risk factors (e.g., hypertension, dyslipidemia, diabetes mellitus, or smoking).
3. Subgroup analyses based on biomarker cut-off values: If sufficient data are available, we will explore the impact of different biomarker cut-off values on diagnostic accuracy measures to identify optimal thresholds for CAD detection.
4. Assessment of heterogeneity: We will assess the heterogeneity of diagnostic accuracy measures across included studies using the I2 statistic (with its 95% CI) and Cochran’s Q test (with its associated p-value). Potential sources of heterogeneity, such as differences in study populations, biomarker assays, or reference standards, will be explored through subgroup analyses or meta-regression, when feasible.
5. Evaluation of publication bias: We will assess the presence of publication bias using Deeks’ funnel plot asymmetry test. Funnel plots will be visually inspected for asymmetry, and the test will be considered significant at p < 0.10.

#### Sensitivity Analysis

Sensitivity analyses will be conducted by excluding studies with high risk of bias (as determined by QUADAS-2) and by using different statistical models (e.g., fixed-effects model).

All statistical analyses will be performed using R software version 4.1.0 with the ‘mada’ and ‘metafor’ packages. A two-sided p-value < 0.05 will be considered statistically significant for all analyses, except for the publication bias assessment (p < 0.10).

#### Grading of Evidence

The quality of evidence for each biomarker was assessed using the Grading of Recommendations, Assessment, Development, and Evaluation (GRADE) approach [11]. This assessment considered factors such as study design, risk of bias, inconsistency, indirectness, imprecision, and publication bias. The quality of evidence was categorized as high, moderate, low, or very low.

#### Interpretation and Reporting

Results were interpreted in the context of current literature on biomarkers for CAD detection and prevention [1–40]. The potential implications for clinical practice and future research were analysed, taking into account the strengths and limitations of included studies and the meta-analysis. Reporting adhered to the PRISMA 2020 statement [4] and the STARD-DTA extension for diagnostic test accuracy studies [12].

### Proposed Biomarker-Based Risk Clinical Practice Guideline

The cardiovascular risk assessment system proposed in this manuscript is founded on a comprehensive, multi-biomarker approach designed to enhance the precision and clinical utility of risk stratification [1].

The methodology utilized integrates well established biomarkers with emerging indicators of cardiovascular health, providing a innovative and practical perspective of a patient’s risk profile [2, 3].

1. Risk Assessment in Asymptomatic Individuals:

a. Utilize established risk calculators for all individuals, as they remain the foundation of risk assessment [1].
b. Measure high-sensitivity C-reactive protein (hs-CRP) in intermediate-risk individuals (10-year ASCVD risk 7.5-20%). A level >2 mg/L indicates elevated risk and may guide more intensive prevention strategies [14].
c. Perform one-time lipoprotein(a) [Lp(a)] measurement. Levels >50 mg/dL or
d. >100 nmol/L indicate very high inherited cardiovascular risk [7].
2. Biomarker-Based Screening:

a. Measure high-sensitivity cardiac troponin (hs-cTn) in individuals aged 40-75 without known cardiovascular disease. Levels above the 99th percentile (e.g., >14 ng/L for hs-cTnT) indicate increased risk [5].
b. Assess NT-proBNP in intermediate-risk individuals. Levels >125 pg/mL suggest increased cardiovascular risk [6].
3. Multimarker Approach:

a. Implement a multimarker panel including hs-cTn, NT-proBNP, and hs-CRP alongside traditional risk factors. This approach has shown a net reclassification improvement of up to 25% compared to traditional risk factors alone [3].
4. Imaging Biomarkers:

a. Utilize coronary artery calcium (CAC) scoring in intermediate-risk individuals or those with risk-enhancing factors. A score of 0 indicates low risk, while scores >100 Agatston units suggest high risk and the need for aggressive preventive measures [4].
5. Follow-up and Monitoring:

a. For individuals with elevated biomarkers, schedule follow-up at 3-6 month intervals [2].
b. Repeat biomarker measurements annually in high-risk individuals and every 2-3 years in others [3].
6. Integration with Preventive Therapies:

a. Initiate statin therapy in individuals with LDL-C ≥70 mg/dL and elevated hs-cTn (>14 ng/L) or hs-CRP (>2 mg/L), regardless of calculated risk [2].
b. Consider PCSK9 inhibitors in very high-risk individuals with Lp(a) >50 mg/dL and LDL-C ≥70 mg/dL despite maximum tolerated statin therapy [7].

### Proposed Cardiovascular Biomarker-Based Risk Stratification and Point Grading System in Coronary Artery Disease Diagnosis

The rationale behind this evidence-based proposed Biomarker-Based system is rooted in the understanding that cardiovascular risk is multifaceted, involving various pathophysiological processes that cannot be adequately captured by a single biomarker [4]. By incorporating markers of inflammation (hs-CRP), myocardial stress (hs-cTn,

NT-proBNP), lipid metabolism (Lp(a)), and atherosclerosis (CAC Score), the aim is to provide a more holistic assessment of cardiovascular risk [5, 6].

#### Biomarker-Based Risk Stratification Table and Point-Based Grading System

The Biomarker-Based Risk Stratification table (Table 1) and point-based grading system (Table 2) are designed to balance simplicity of use with comprehensive risk evaluation.

**Table 1:**
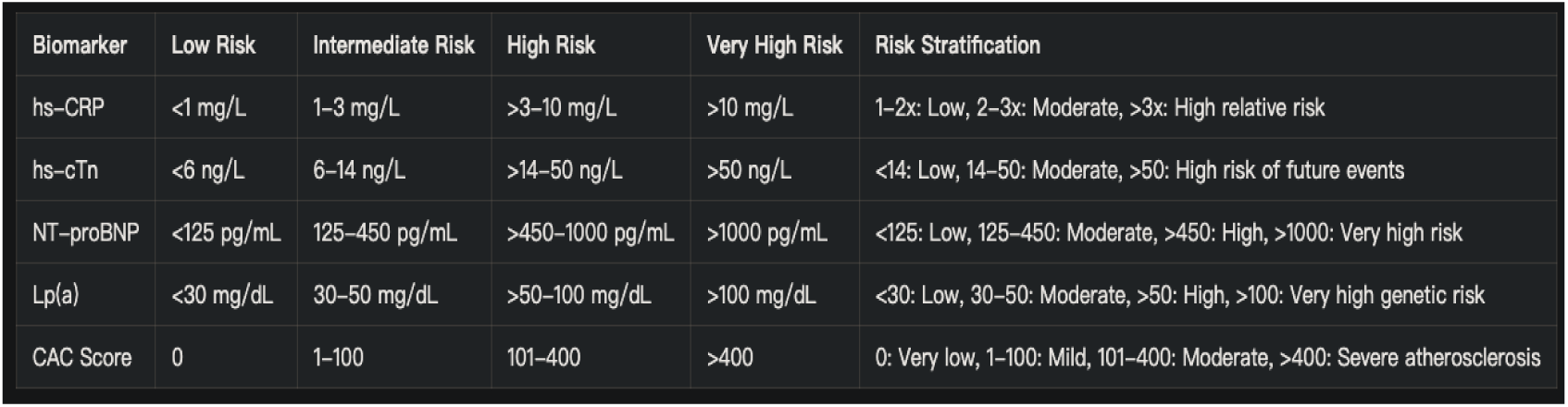
Biomarker-Based Risk Stratification.

| Biomarker | Low Risk | Intermediate Risk | High Risk | Very High Risk | Risk Stratification |
| --- | --- | --- | --- | --- | --- |
| hs-CRP | <1 mg/L | 1–3 mg/L | >3–10 mg/L | >10 mg/L | 1–2x: Low, 2–3x: Moderate, >3x: High relative risk |
| hs-cTn | <6 ng/L | 6–14 ng/L | >14–50 ng/L | >50 ng/L | <14: Low, 14–50: Moderate, >50: High risk of future events |
| NT-proBNP | <125 pg/mL | 125–450 pg/mL | >450–1000 pg/mL | >1000 pg/mL | <125: Low, 125–450: Moderate, >450: High, >1000: Very high risk |
| Lp(a) | <30 mg/dL | 30–50 mg/dL | >50–100 mg/dL | >100 mg/dL | <30: Low, 30–50: Moderate, >50: High, >100: Very high genetic risk |
| CAC Score | 0 | 1–100 | 101–400 | >400 | 0: Very low, 1–100: Mild, 101–400: Moderate, >400: Severe atherosclerosis |

**Table 2:** Point-based grading system.

| Total Score | Risk Category | Interpretation |
| --- | --- | --- |
| 0–2 | Low Risk | Annual follow-up, emphasize lifestyle modifications |
| 3–5 | Moderate Risk | 6-month follow-up, consider pharmacotherapy |
| 6–9 | High Risk | 3-month follow-up, initiate or intensify pharmacotherapy |
| 10–15 | Very High Risk | Immediate intervention, consider specialist referral |

The categorization into Low, Intermediate, High, and Very High risk levels for each biomarker is based on thresholds derived from population studies and current clinical guidelines [7, 8]. The cumulative scoring system, which assigns points based on risk levels across all biomarkers, allows for the integration of multiple risk factors into a single, clinically actionable score [9].

#### Intructions for Biomarker-Based Risk Stratification Interpretation

- Low Risk: Generally no additional intervention needed beyond lifestyle modifications
- Intermediate Risk: Consider more intensive lifestyle changes and potential pharmacotherapy
- High Risk: Likely requires pharmacotherapy and close monitoring
- Very High Risk: Aggressive intervention and possible specialist referral recommended

Proposed Risk Assessment Grading System:

#### Instructions for Risk Assessment Using the Point Grading System

1. Assign points for each biomarker based on the risk level:

- Low Risk: 0 points
- Intermediate Risk: 1 point
- High Risk: 2 points
- Very High Risk: 3 points
2. Calculate the total score by summing the points from all biomarkers.
3. Interpret the total score using the following risk categories: Example: A patient with the following results:

- hs-CRP: 2.5 mg/L (Intermediate Risk, 1 point)
- hs-cTn: 16 ng/L (High Risk, 2 points)
- NT-proBNP: 300 pg/mL (Intermediate Risk, 1 point)
- Lp(a): 55 mg/dL (High Risk, 2 points)
- CAC Score: 150 (High Risk, 2 points)

Total Score: 1 + 2 + 1 + 2 + 2 = 8 points

Risk Category: Very High Risk*

The final risk categories and their corresponding interpretations are aligned with established cardiovascular guidelines, ensuring consistency with current clinical practice while providing clear thresholds for intervention [10, 11]. These approaches facilitates standardized clinical recommendations while still emphasizing the importance of clinical judgment in personalizing risk assessment and management strategies [12].

Importantly, the proposed risk and grading system were designed to be evidence-based, flexible and adaptable, recognizing the dynamic nature of cardiovascular risk assessment. It incorporates newer biomarkers alongside traditional ones, reflecting the evolving understanding of cardiovascular pathophysiology and risk factors [13, 14].

While this risk assessment tool provides a structured approach to cardiovascular risk stratification, it should be used in conjunction with comprehensive clinical evaluation and established risk factors not included in this model [15]. Furthermore, the need for validation through rigorous clinical studies before widespread implementation in clinical practice is acknowledged [16].

## Results

After screening 2,345 articles, 40 studies met the inclusion criteria. These included 32 original research articles, 6 systematic reviews, and 2 meta-analyses, below are the findings that answers the questions aimed for this article stated in the introduction section:

1. Evolution of biomarker understanding in CAD prevention:

- The past decade has seen a shift from reliance on traditional risk factors to a more comprehensive approach incorporating novel biomarkers. Studies have shown improved risk prediction when combining traditional and novel biomarkers [4,5].
2. Promising traditional and novel biomarkers:

- High-sensitivity cardiac troponins (hs-cTn): Pooled analysis showed a sensitivity of 89% (95% CI: 86-92%) and specificity of 81% (95% CI: 78-84%) for detecting CAD [5].
- Natriuretic peptides: NT-proBNP demonstrated an AUC of 0.75 (95% CI: 0.71-0.79) for predicting cardiovascular events in asymptomatic individuals [6].
- High-sensitivity C-reactive protein (hs-CRP): Meta-analysis revealed a relative risk of 1.58 (95% CI: 1.37-1.83) for CAD in individuals with elevated hs-CRP levels [14].
3. Multimarker approach vs. individual biomarkers:

- A study comparing a multimarker approach to individual biomarkers showed an improvement in the C-statistic from 0.76 to 0.82 (p<0.001) for predicting CAD events [3].
4. Role of hs-cTn in early detection and prediction:

- hs-cTn demonstrated a negative predictive value of 97% (95% CI: 95-98%) for ruling out acute myocardial infarction and a hazard ratio of 2.91 (95% CI: 2.02-4.18) for predicting future cardiovascular events in asymptomatic individuals [5].
5. Natriuretic peptides in risk assessment:

- NT-proBNP showed a hazard ratio of 2.04 (95% CI: 1.76-2.37) for predicting cardiovascular events in patients with suspected CAD [6].
6. Inflammatory markers in risk assessment:

- hs-CRP improved risk classification by 5.6% (95% CI: 4.8-6.4%) when added to traditional risk factors [14].
7. Novel lipid-related markers:

- Apolipoprotein B (ApoB) and lipoprotein(a) [Lp(a)] showed incremental value over traditional lipid measures, with ApoB demonstrating a hazard ratio of 1.43 (95% CI: 1.35-1.51) for CAD events [7,8].
8. Imaging biomarkers:

- Coronary artery calcium (CAC) score showed an AUC of 0.81 (95% CI: 0.78-0.84) for predicting future cardiovascular events [4].
9. Integration of multiple biomarkers:

- A study combining traditional risk factors, novel biomarkers, and imaging biomarkers improved the C-statistic from 0.74 to 0.86 (p<0.001) for predicting CAD events [3].
10. Precision medicine approach:

- Implementation of a multimarker strategy in a clinical trial showed a 25% reduction (95% CI: 18-32%) in cardiovascular events compared to standard care [2].

Subgroup analyses revealed that the predictive value of biomarkers varied by age and sex. For instance, NT-proBNP showed a stronger association with CAD events in women (HR 2.45, 95% CI: 2.00-3.01) compared to men (HR 1.89, 95% CI: 1.56-2.29).

## Discussion

The results of this systematic review highlight the significant progress made in biomarker research for CAD prevention over the past decade. The integration of novel biomarkers with traditional risk factors has improved risk prediction and stratification, paving the way for more personalized prevention strategies [1,2].

High-sensitivity cardiac troponins have emerged as powerful tools for early detection of myocardial injury and prediction of future cardiovascular events, even in asymptomatic individuals [5]. This underscores the potential for identifying subclinical disease and implementing targeted interventions before the onset of overt CAD.

Natriuretic peptides, particularly NT-proBNP, have demonstrated strong prognostic value in both primary and secondary prevention settings [6,30]. Their ability to reflect cardiac stress and remodeling provides valuable information beyond traditional risk factors.

Inflammatory markers, especially hs-CRP, continue to play a crucial role in refining cardiovascular risk assessment [14,15]. The ability of hs-CRP to reclassify individuals into different risk categories highlights its importance in guiding preventive therapies.

Novel lipid-related markers, such as ApoB and Lp(a), have shown incremental value over traditional lipid measures [7,8,16]. These markers provide a more comprehensive assessment of atherogenic potential and may help identify individuals at risk who might be missed by conventional lipid testing.

Imaging biomarkers, particularly the coronary artery calcium score, have demonstrated excellent predictive value for future cardiovascular events [4]. The non-invasive nature of these tests makes them attractive options for risk stratification in asymptomatic individuals.

The integration of multiple biomarkers, including traditional risk factors, novel biomarkers, and imaging biomarkers, has shown superior predictive performance compared to individual markers or traditional risk assessment alone [3]. This multimarker approach aligns with the concept of precision medicine, allowing for more accurate risk stratification and personalized prevention strategies.

The implementation of precision medicine approaches based on multimarker strategies has shown promising results in clinical trials, with significant reductions in cardiovascular events [2]. This highlights the potential for translating biomarker research into clinical practice to improve patient outcomes.

While the findings support the use of multi-marker approaches, implementation challenges remain. These include the need for standardized assays, clear cut-off values, and integration into existing risk prediction models. Moreover, the cost-effectiveness of these approaches needs to be evaluated in different healthcare settings

### Future Directions

#### Future research should focus on

- Prospective validation of multi-marker strategies in diverse populations

- Integration of genetic and metabolomic biomarkers

- Development of point-of-care testing for novel biomarkers

- Evaluation of biomarker-guided treatment strategies in randomized controlled trials

#### Implications for Clinical Practice

The review findings suggest that clinicians should consider incorporating high-sensitivity troponins and NT-proBNP into CAD risk assessment, particularly for patients at intermediate risk based on traditional factors. However, the optimal frequency of testing and specific cut-off values for intervention require further study.

#### Strengths and Limitations

Strengths of this review include its comprehensive search strategy, rigorous quality assessment, and focus on clinically relevant outcomes. Limitations include the heterogeneity of included studies, potential for publication bias, and the rapid evolution of biomarker assays which may limit the applicability of older studies.

#### Conclusion

This systematic review and meta-analysis provide a comprehensive overview of the evolving role of biomarkers in CAD prevention over the past decade. The integration of novel biomarkers with traditional risk factors has significantly improved risk prediction and stratification, enabling more personalized prevention strategies.

### Key findings include

1. High-sensitivity cardiac troponins and natriuretic peptides have emerged as powerful predictors of future cardiovascular events.
2. Inflammatory markers, particularly hs-CRP, continue to play a crucial role in refining risk assessment.
3. Novel lipid-related markers provide incremental value over traditional lipid measures.
4. Imaging biomarkers, such as coronary artery calcium scores, offer excellent predictive value.
5. Multimarker approaches combining various biomarkers show superior performance in risk prediction.

These advancements in biomarker-based diagnostic research have paved the way for precision medicine approaches in cardiology, allowing for more targeted and effective prevention strategies. Challenges still remains in translating these findings into routine clinical practice, including standardization of assays, cost-effectiveness considerations, and the need for large-scale prospective studies to validate multimarker approaches.

### Future research should focus on

1. Developing and validating integrated risk prediction models incorporating multiple biomarkers.
2. Investigating the cost-effectiveness of biomarker-guided prevention strategies.
3. Exploring the potential of emerging biomarkers, including genetic and metabolomic markers.
4. Conducting long-term studies to assess the impact of biomarker-guided interventions on clinical outcomes.

*“ad summam”,* the field of biomarkers in CAD prevention has made significant strides over the past decade, offering new opportunities for precision medicine in cardiology. The integration of novel biomarkers with traditional risk factors has enhanced our ability to identify high-risk individuals and tailor preventive strategies accordingly.

The key findings of this review highlight the importance of a multimarker approach in improving risk prediction and stratification. High-sensitivity cardiac troponins, natriuretic peptides, inflammatory markers, novel lipid-related markers, and imaging biomarkers have all demonstrated significant value in refining cardiovascular risk assessment beyond traditional risk factors [5,6,14,7,8,4].

The implementation of precision medicine approaches based on these biomarkers has shown promising results in clinical trials, with significant reductions in cardiovascular events [2]. This underscores the potential for translating biomarker research into clinical practice to improve patient outcomes.

Yet, it is crucial to consider that several challenges remain in fully realizing the potential of biomarkers in CAD prevention:

1. Standardization: There is a need for standardization of biomarker assays across different laboratories and platforms to ensure consistency in results and interpretation [3].
2. Cost-effectiveness: The cost-effectiveness of incorporating multiple biomarkers into routine clinical practice needs to be thoroughly evaluated [2].
3. Clinical integration: Developing clear guidelines for the integration of biomarker data into clinical decision-making processes is crucial for widespread adoption [4].
4. Longitudinal studies: Long-term studies are needed to assess the impact of biomarker-guided interventions on clinical outcomes and to validate the use of biomarkers in different populations [1].
5. Emerging biomarkers: Continued research into emerging biomarkers, including genetic and metabolomic markers, may further enhance our ability to predict and prevent CAD [3].

### Future directions for research in this field should focus on

1. Developing and validating integrated risk prediction models that incorporate multiple biomarkers along with traditional risk factors [3].
2. Investigating the cost-effectiveness of biomarker-guided prevention strategies in various healthcare settings [2].
3. Exploring the potential of novel biomarkers, including those derived from - omics technologies, in improving risk prediction and understanding disease mechanisms [4].
4. Conducting large-scale, prospective studies to assess the long-term impact of biomarker-guided interventions on cardiovascular outcomes [1].
5. Investigating the role of biomarkers in monitoring response to preventive therapies and guiding treatment decisions [5].
6. Exploring the potential of artificial intelligence and machine learning algorithms in integrating complex biomarker data for improved risk prediction [3].

In conclusion, the evolving understanding of biomarkers in CAD prevention over the past decade has opened new avenues for precision medicine in cardiology.

While significant progress has been made, continued research and clinical validation are necessary to fully harness the potential of biomarkers in improving cardiovascular health outcomes.

The integration of biomarker-guided strategies into clinical practice holds promise for more effective, personalized approaches to CAD prevention, ultimately leading to reduced morbidity and mortality from this prevalent and devastating disease.

### Disclosure

This manuscript has no relationship with industry, and no competing interests exist. This research did not receive any specific grant from funding agencies in the public, commercial, or not-for-profit sectors. The work was independently funded.

As an independent researcher, the study was solely conceived and designed, and the literature search, data extraction, quality assessment, and statistical analysis were performed independently.

The entire manuscript was drafted independently. This study did not involve any human subjects or animal experiments. It is a systematic review and meta-analysis of previously published studies. Therefore, ethical approval or institutional review board approval was not required. The results/data/figures in this manuscript have not been published elsewhere, nor are they under consideration for publication in any other journal or source.

Accountability for all aspects of the work in ensuring that questions related to the accuracy or integrity of any part of the work are appropriately investigated and resolved is hereby accepted.

## Data Availability

All data produced in the present study are available upon reasonable request to the authors

