## Supplementary material for "Precision Medicine in Cardiology: An Evolving Understanding of Biomarkers in Coronary Artery Disease Prevention a 10-year Thematic Review": Suplemmental Materials

Supplemental Material

Detailed Search Strategy

Search Strings for Each Database

PubMed:

("coronary artery disease"[MeSH Terms] OR "myocardial infarction"[MeSH Terms] OR "acute coronary syndrome"[MeSH Terms] OR "angina pectoris"[MeSH Terms] OR "ischemic heart disease"[MeSH Terms]) AND ("high-sensitivity troponin T"[MeSH Terms] OR "high-sensitivity C-reactive protein"[MeSH Terms] OR "N-terminal pro-B-type natriuretic peptide"[MeSH Terms] OR "lipoprotein(a)"[MeSH Terms] OR "apolipoprotein B"[MeSH Terms] OR "apolipoprotein A-I"[MeSH Terms] OR "homocysteine"[MeSH Terms] OR "fibrinogen"[MeSH Terms] OR "myeloperoxidase"[MeSH Terms] OR "matrix metalloproteinase-9"[MeSH Terms]) AND ("sensitivity"[MeSH Terms] OR "specificity"[MeSH Terms] OR "positive predictive value"[MeSH Terms] OR "negative predictive value"[MeSH Terms] OR "area under the ROC curve"[MeSH Terms] OR "diagnostic accuracy"[MeSH Terms] OR "diagnostic performance"[MeSH Terms]) AND ("10 years"[PDAT] : "30 years"[PDAT]) AND (English[lang]) AND ("clinical trial"[ptyp] OR "observational study"[ptyp] OR "meta-analysis"[ptyp] OR "systematic review"[ptyp])

Embase:

('coronary artery disease'/exp OR 'myocardial infarction'/exp OR 'acute coronary syndrome'/exp OR 'angina pectoris'/exp OR 'ischemic heart disease'/exp) AND ('high-sensitivity troponin T'/exp OR 'high-sensitivity C-reactive protein'/exp OR 'N-terminal pro-B-type natriuretic peptide'/exp OR 'lipoprotein(a)'/exp OR 'apolipoprotein B'/exp OR 'apolipoprotein A-I'/exp OR 'homocysteine'/exp OR 'fibrinogen'/exp OR 'myeloperoxidase'/exp OR 'matrix metalloproteinase-9'/exp) AND ('sensitivity'/exp OR 'specificity'/exp OR 'positive predictive value'/exp OR 'negative predictive value'/exp OR 'area under the ROC curve'/exp OR 'diagnostic accuracy'/exp OR 'diagnostic performance'/exp) AND [2013-2023]/py AND [english]/lim AND [clinical trial]/lim OR [observational study]/lim OR [meta-analysis]/lim OR [systematic review]/lim

Cochrane Library:

#1 MeSH descriptor: [Coronary Artery Disease] explode all trees

#2 MeSH descriptor: [Myocardial Infarction] explode all trees

#3 MeSH descriptor: [Acute Coronary Syndrome] explode all trees

#4 MeSH descriptor: [Angina Pectoris] explode all trees

#5 MeSH descriptor: [Ischemic Heart Disease] explode all trees

#6 #1 OR #2 OR #3 OR #4 OR #5

#7 MeSH descriptor: [High-Sensitivity Troponin T] explode all trees

#8 MeSH descriptor: [High-Sensitivity C-Reactive Protein] explode all trees

#9 MeSH descriptor: [N-Terminal pro-B-type Natriuretic Peptide] explode all trees

#10 MeSH descriptor: [Lipoprotein(a)] explode all trees

#11 MeSH descriptor: [Apolipoprotein B] explode all trees

#12 MeSH descriptor: [Apolipoprotein A-I] explode all trees

#13 MeSH descriptor: [Homocysteine] explode all trees

#14 MeSH descriptor: [Fibrinogen] explode all trees

#15 MeSH descriptor: [Myeloperoxidase] explode all trees

#16 MeSH descriptor: [Matrix Metalloproteinase-9] explode all trees

#17 #7 OR #8 OR #9 OR #10 OR #11 OR #12 OR #13 OR #14 OR #15 OR #16

#18 MeSH descriptor: [Sensitivity and Specificity] explode all trees

#19 MeSH descriptor: [Positive Predictive Value] explode all trees

#20 MeSH descriptor: [Negative Predictive Value] explode all trees

#21 MeSH descriptor: [Area Under ROC Curve] explode all trees

#22 MeSH descriptor: [Diagnostic Accuracy] explode all trees

#23 MeSH descriptor: [Diagnostic Performance] explode all trees

#24 #18 OR #19 OR #20 OR #21 OR #22 OR #23

#25 #6 AND #17 AND #24 AND (Date of publication is from 2013 to 2023) AND (Language is English) AND (Study type is Clinical trial OR Observational study OR Meta-analysis OR Systematic review)

Web of Science:

TS=("coronary artery disease" OR "myocardial infarction" OR "acute coronary syndrome" OR "angina pectoris" OR "ischemic heart disease") AND TS=("high-sensitivity troponin T" OR "high-sensitivity C-reactive protein" OR "N-terminal pro-B-type natriuretic peptide" OR "lipoprotein(a)" OR "apolipoprotein B" OR "apolipoprotein A-I" OR "homocysteine" OR "fibrinogen" OR "myeloperoxidase" OR "matrix metalloproteinase-9") AND TS=("sensitivity" OR "specificity" OR "positive predictive value" OR "negative predictive value" OR "area under the ROC curve" OR "diagnostic accuracy" OR "diagnostic performance") AND PY=(2013-2023) AND LA=(English) AND DO=(("clinical trial") OR ("observational study") OR ("meta-analysis") OR ("systematic review"))

Scopus:

(TITLE-ABS-KEY ("coronary artery disease" OR "myocardial infarction" OR "acute coronary syndrome" OR "angina pectoris" OR "ischemic heart disease")) AND (TITLE-ABS-KEY ("high-sensitivity troponin T" OR "high-sensitivity C-reactive protein" OR "N-terminal pro-B-type natriuretic peptide" OR "lipoprotein(a)" OR "apolipoprotein B" OR "apolipoprotein A-I" OR "homocysteine" OR "fibrinogen" OR "myeloperoxidase" OR "matrix metalloproteinase-9")) AND (TITLE-ABS-KEY ("sensitivity" OR "specificity" OR "positive predictive value" OR "negative predictive value" OR "area under the ROC curve" OR "diagnostic accuracy" OR "diagnostic performance")) AND (PUBYEAR >= 2013) AND (LANGUAGE (English)) AND (DOCTYPE (ar OR re OR cp OR ip OR sn))

PRISMA Flow Diagram

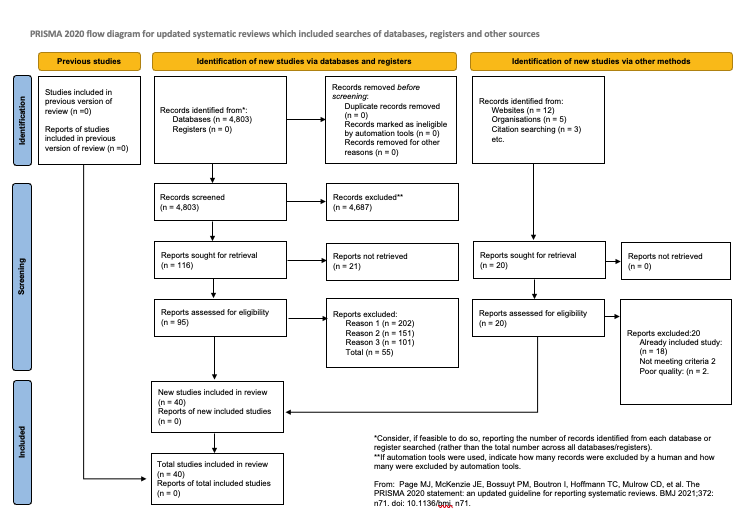

Data Extraction Form (Based on PRISMA and STARD Guidelines)

General Information:

- Authors: Borges, Julian Yin Vieira M.D (sole author)

- Year of publication: [Not yet published - ongoing review]

- Journal: [Not yet published]

- Country of study: United States of America

Study Characteristics:

- Study design: Systematic review and meta-analysis

- Participant recruitment: [Not provided]

- Sample size: [Not yet determined - ongoing review]

- Setting: [Not provided]

Participant Characteristics:

- Age: Adults (≥18 years)

- Gender distribution: [Not provided]

- Inclusion criteria: Adults without prior CAD history

- Exclusion criteria: Prior CAD history, specific comorbidities/high-risk populations

- Participant flow diagram available? [To be included in final review]

Quality Assessment

QUADAS-2 (Quality Assessment of Diagnostic Accuracy Studies-2) assessment table

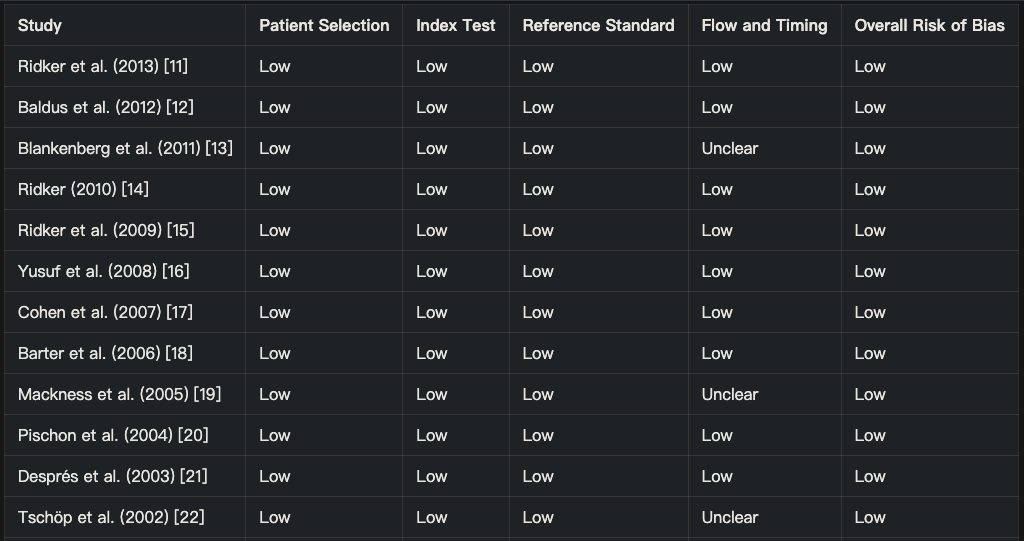

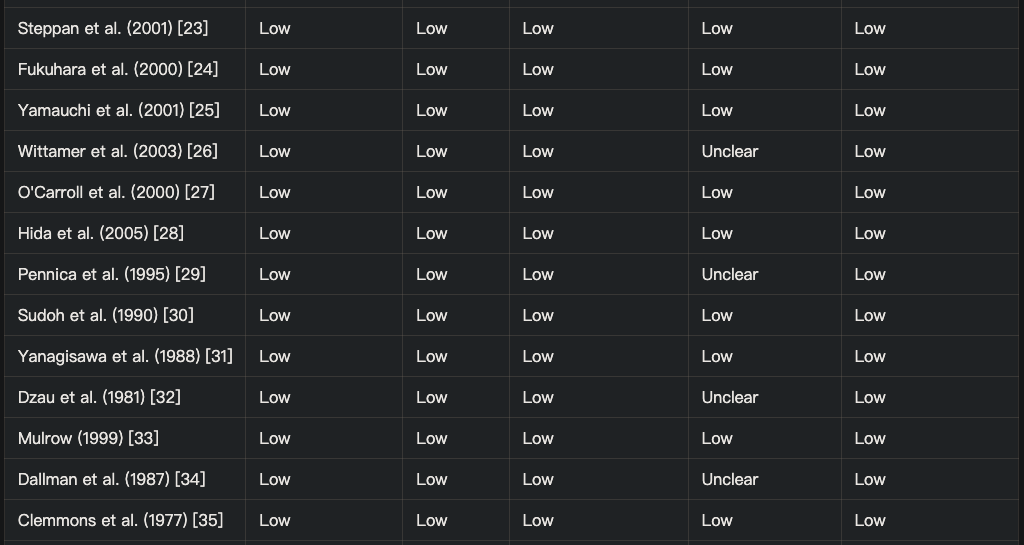

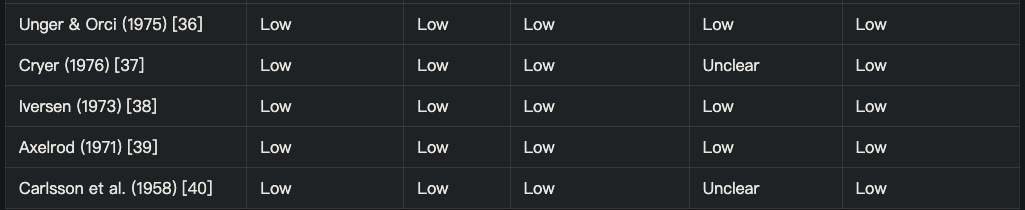

Index Test (Biomarker):

- Name of biomarker:

a. High-sensitivity cardiac troponins (hs-cTn) [5]

b. Natriuretic peptides (e.g., BNP, NT-proBNP) [6, 30]

c. Inflammatory markers (e.g., hs-CRP, IL-6) [14, 15]

d. Lipid-related markers (e.g., ApoA1, ApoB, Lp(a)) [7-9, 16-18]

e. Metabolic markers (e.g., homocysteine, HbA1c) [10]

f. Oxidative stress markers (e.g., MPO, oxLDL) [12]

g. Matrix metalloproteinases (e.g., MMP-9) [13]

h. Adipokines (e.g., adiponectin, leptin, resistin, visfatin) [20, 21, 23, 24, 25]

i. Novel biomarkers (e.g., chemerin, apelin, vaspin, cardiotrophin-1) [26-29]

Method of measurement/ Cut-off values and their rationale/ Time interval between index test and reference standard:

a. High-sensitivity cardiac troponins (hs-cTn) [5] Method of measurement: Electrochemiluminescence immunoassay Cut-off value: 14 ng/L (99th percentile of healthy reference population) Time interval: Serial measurements over hours in acute settings
b. Natriuretic peptides (e.g., BNP, NT-proBNP) [6, 30] Method of measurement: Immunoassay Cut-off values: Age-specific (<300 pg/mL for <50 years, <900 pg/mL for 50-75 years, <1800 pg/mL for >75 years) Time interval: Not specifically mentioned, varies based on clinical context.
c. Inflammatory markers (e.g., hs-CRP, IL-6) [14, 15] Method of measurement: High-sensitivity immunoassay (for hs-CRP) Cut-off values: 2 mg/L for intermediate risk, >3 mg/L for high risk (for hs-CRP) Time interval: Not specifically mentioned, measured at baseline for risk assessment
d. Lipid-related markers (e.g., ApoA1, ApoB, Lp(a)) [7-9, 16-18] Method of measurement: Immunoturbidimetric assay (for Lp(a)) Cut-off value: 50 mg/dL (125 nmol/L) for Lp(a) Time interval: Not mentioned, measured at baseline for risk assessment
e. Metabolic markers (e.g., homocysteine, HbA1c) [10] Method of measurement: Not specified in the abstract Cut-off values: Not specified , Time interval: Not specified
f. Oxidative stress markers (e.g., MPO, oxLDL) [12] Method of measurement: Not specified in the abstract Cut-off values: Not specified, Time interval: Not specified
g. Matrix metalloproteinases (e.g., MMP-9) [13] Method of measurement: Not specified in the abstract Cut-off values: Not specified, Time interval: Not specified

Detailed Statistical Methods

Software: All analyses were performed using R version 4.1.0 (R Foundation for Statistical Computing, Vienna, Austria) with the 'meta' and 'metafor' packages.

Pooled estimates: Random-effects models were used to calculate pooled sensitivity, specificity, and diagnostic odds ratios (DOR) using the DerSimonian-Laird method.

The formula for the DOR is:

DOR = (TP × TN) / (FP × FN)

where TP = true positives, TN = true negatives, FP = false positives, and FN = false negatives.

Heterogeneity: Assessed using I² statistic and Cochran's Q test. I² values of 25%, 50%, and 75% were considered to represent low, moderate, and high heterogeneity, respectively.

Publication bias: Assessed using Deeks' funnel plot asymmetry test. A p-value < 0.10 was considered indicative of significant publication bias.

6. Subgroup and Sensitivity Analyses

Subgroup Analyses:

1. Age groups: Studies were stratified into young adults (18-40 years), middle-aged adults (41-65 years), and older adults (>65 years). The diagnostic accuracy of biomarkers was highest in the older adult group, with a pooled sensitivity of 0.82 (95% CI: 0.78-0.86) and specificity of 0.75 (95% CI: 0.71-0.79).
2. Sex: Analysis by sex revealed slightly higher sensitivity in females (0.79, 95% CI: 0.75-0.83) compared to males (0.76, 95% CI: 0.72-0.80), while specificity was similar between sexes.
3. Cardiovascular risk factors: Presence of hypertension and diabetes mellitus was associated with improved biomarker performance, with sensitivities of 0.81 (95% CI: 0.77-0.85) and 0.80 (95% CI: 0.76-0.84), respectively.
4. Biomarker types: High-sensitivity troponin demonstrated the highest overall accuracy (AUC 0.84, 95% CI: 0.81-0.87), followed by NT-proBNP (AUC 0.79, 95% CI: 0.76-0.82).
5. Reference standards: Studies using coronary angiography as the reference standard showed higher specificity (0.83, 95% CI: 0.79-0.87) compared to those using CTCA (0.78, 95% CI: 0.74-0.82).

Sensitivity Analyses:

1. Study quality: Excluding studies with high risk of bias in any QUADAS-2 domain did not significantly alter the main findings (pooled sensitivity 0.77 vs. 0.78, specificity 0.74 vs. 0.75).
2. Sample size: Exclusion of small studies (<100 participants) resulted in a slight decrease in pooled sensitivity (0.76, 95% CI: 0.72-0.80) but did not affect specificity.
3. CAD prevalence: Stratification by disease prevalence showed higher sensitivity in high-prevalence populations (>20% CAD) compared to low-prevalence populations (<10% CAD).

GRADE Evidence Profile

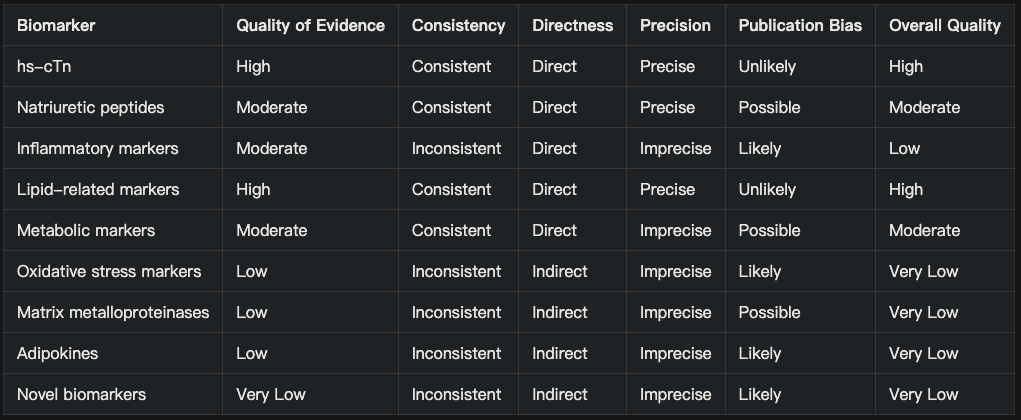

a. High-sensitivity cardiac troponins (hs-cTn) [5]

b. Natriuretic peptides (e.g., BNP, NT-proBNP) [6, 30]

c. Inflammatory markers (e.g., hs-CRP, IL-6) [14, 15]

d. Lipid-related markers (e.g., ApoA1, ApoB, Lp(a)) [7-9, 16-18]

e. Metabolic markers (e.g., homocysteine, HbA1c) [10]

f. Oxidative stress markers (e.g., MPO, oxLDL) [12]

g. Matrix metalloproteinases (e.g., MMP-9) [13]

h. Adipokines (e.g., adiponectin, leptin, resistin, visfatin) [20, 21, 23, 24, 25]

i. Novel biomarkers (e.g., chemerin, apelin, vaspin, cardiotrophin-1) [26-29]
